# Voting Patterns, Mortality, and Health Inequalities in England: A replication and extension of Smith and Dorling (1996)

**DOI:** 10.1101/2024.06.26.24309517

**Authors:** Charles Rahal, Josh Knight, Veline L’Esperance, Melinda C. Mills, Philip M. Clarke

**Author notes:** **For Correspondence:** Professor Philip Clarke, Health Economics Research Centre, Nuffield Department of Population Health, Old Road Campus, OX3 7LF, University of Oxford, United Kingdom. **Contributors:** PMC led on the initial conception and drafting of the manuscript. CR lead on all data analysis, visualisation, and replication materials. All authors contributed to the revision of the manuscript.

## Abstract

**Objectives:** To investigate the relationship between voting patterns, mortality, health, and disability across England, replicating and extending a BMJ study from 1996.

**Design:** Observational study using data from the Office of National Statistics and the House of Commons Library, mapped to electoral constituencies.

**Setting:** England, UK.

**Participants:** The mortality, health and disability data come from the English population across multiple publicly available datasets and are cross tabulated against information on voting in the 2019 and 2024 UK General Election in constituencies in England.

**Main Outcome Measures:** Age-standardised mortality rate (ASMR) for 2021, as well as Health and Disability metrics from the UK Census of 2021.

**Results:** When observing the proportion of vote-share for Labour at the constituency level in both elections, there was a strong, positive correlation with ASMR. In the 2019 election, this was *r*=0.708 for males, and *r*=0.653 for females. For the 2024 election, this was *r*=0.540 for males, and *r*=0.539 for females. There are also correlations between Labour vote shares and measures of health, but far less substantially for measures of disability. The strongest correlations were almost unilaterally observed against the proportion of votes cast in a constituency. A marked deviation was in the 2019 election where there was also a small, but positive correlation with voting Conservative and poor health (*r*=0.035) and disability (*r*=0.081), but not for ASMR (*r*=-0.489 for females, *r*=-0545 for males). Strong, positive correlations were also observed between all covariates and vote share for the Brexit Party (2019) and Reform UK (2024).

**Conclusions:** Health and mortality inequalities across England remain high, with trends largely following previous political patterning. People needing to rely on state provisions likely vote for the political party they believe will be best placed to solve health and structural issues.

## 1 Introduction

Health and mortality inequalities across England have been significant and persistent, with consistent associations found between poor health and higher mortality in areas of socioeconomic deprivation and limited access to health services, along with demographic variation.[1, 2] Since the publication of the Acheson Report in 1998,[3] there have been more than twenty subsequent reports and inquiries into health inequalities. These document the large variation in health and mortality across England and propose a wide range of public health and related strategies to reduce structural inequalities. Several contained specific targets, such as the Labour Government’s ambition in 2001 to “reduce by at least 10 per cent the gap between the quintile of areas with the lowest life expectancy at birth and the population as a whole”,[4] with some evidence that this goal was achieved.[5] In 2022, the Conservative Government’s Levelling Up agenda had an explicit mission that “by 2030, the gap in Healthy Life Expectancy between local areas where it is highest and lowest will have narrowed, and by 2035 healthy life expectancy will rise by five years”.[6] In the run up to the 2024 election, the then Labour party committed itself to “improve healthy life expectancy for all and halve the gap in healthy life expectancy between different regions of England”.[7]

Previous research used classic measures such as deprivation indices or health care access to explain geographical variations in health and mortality. A less explored issue is how these relate to voting behaviour. The current study replicates and extends work from the 1990s published in the BMJ, exploring the relationship between mortality rates, deprivation, and voting patterns in English and Welsh constituencies during the three electoral cycles of 1983, 1987, and 1992.[8] The study found a strong, statistically significant bivariate relationship between voting patterns and standardised mortality ratios, a measure of relative mortality. Proportions of Conservative electoral votes within a constituency were negatively correlated (1983, –0.76; 1987, –0.74; 1992 -0.74) with higher mortality; conversely, there were large and significant positive correlations (0.76; 0.77; 0.73) with the proportion of votes won by the Labour Party (all p¡0.0001). After almost three decades after this original publication in BMJ, we revisit the political geography of health and mortality inequalities in England. The new analysis covers a pivotal period in British politics where the 2019 election reinforced Boris Johnson’s Conservative Party majority after a period of political uncertainty. This was then overturned by a large majority in the 2024 election by the Labour Party under the leadership of Kier Starmer, with the Labour Party having been out of power since 2010.

## 2 Data and Methods

This observational study is based on official published statistics of mortality, health, disability, and voting patterns. It contributes a descriptive understanding of how levels of mortality vary across geographically based electoral constituencies in England.

### 2.1 Data

To represent constituency voting patterns, data from the Westminster House of Commons Library for the most recent two (2019 and 2024) elections was analysed based on the proportion of votes cast for major parties across English constituencies. Three measures were included in the analysis: age-standardised mortality rates (ASMR) were used across constituencies (based on 2021 data), as well as information on health and disability from the 2021 UK Census. The ASMR data was age-standardised based on the 2013 European Standard Population and provided as per 100,000 rates by sex.[9] The year of death was based on the date of the registration of the death. From the UK Census of 2021, we operationalise two different metrics, but both in the same way: first, the percentage of people who self-report that they are in a state of ‘Not good health’, and second, the percentage of people who are ‘Disabled under the Equality Act’. Under the 2010 Equality Act in the UK, a person is considered disabled if they have a physical or mental impairment that has a substantial and long-term adverse effect on their ability to carry-out normal day-to-day activities. This can include impairment (illnesses or conditions such as autism, dyslexia), long-term or progressive conditions (Alzheimer’s) or certain conditions even if they do not have a substantial impact on daily activities (cancer, HIV, multiple sclerosis).[10]

Naturally, and while our covariates represent the state of mortality, health and disability in the year of 2021 (i.e., a midpoint between two important elections), a critical electoral boundary change occurred in 2023. There were 533 constituencies contested (in England) in the 2019 election, whereas in 2024 there were changes in most constituencies’ composition, and the number of constituencies changed to 543. The implications of this are that when we link the ONS ASMR data (which has 533 values), we are limited to analysing only 519 constituencies in 2024, given that fourteen constituencies are not provided through a recent ONS lookup file to their most proximal constituencies from 2019. See our online supplementary materials for a further discussion of this. With regards to the observational Census data provided directly by the ONS, we have a full sample of 533 and 543 constituencies in each of the 2019 and 2024 elections. These sample sizes are also reported in Table 1. We primarily analyse Labour voting share in Figures 1-2 (given that they are the most recently successful party in 2024), but we also provide results for other political parties via our online supplementary materials, and in Table 1. Specifically, we include supplementary visualisation between correlates with vote share for the Conservative Party, the Liberal Democrats, and the proportion of people voting. In 2019 and 2024, we analyse results from the Brexit Party and Reform UK, respectively (both in Table 1 and in our online materials). Vote shares are conceptualised as the number of votes for a party, divided by the number of ‘valid votes’ (as opposed to all votes cast, with a small number of votes cast being invalid). The exception is the analysis of what is termed ‘Vote’ in Table 1, which is conceptualised as the sum of valid and invalid votes divided by the total number of people eligible in the constituency. For statistical results, we also drop the constituency of the Speaker of the House, who held the constituency of Chorley for both elections.

**Table 1.** Political party by age-standardised mortality rate and proportion of not good health and disability, stratified by sex and election year. The table shows political party voting behaviour by Age Standardized Mortality Rate (ASMR) for females and males, and the proportion of those in not good health and with disability (Office for National Statistics).

| Political Party | ASMR (f) |  | ASMR (m) |  | Health |  | Disability |  |
| --- | --- | --- | --- | --- | --- | --- | --- | --- |
|  | 2019 | 2024 | 2019 | 2024 | 2019 | 2024 | 2019 | 2024 |
| Conservative | -0.489 | -0.602 | -0.545 | -0.627 | 0.035 | -0.185 | 0.081 | -0.138 |
| Labour | 0.653 | 0.539 | 0.708 | 0.540 | 0.145 | 0.211 | 0.028 | 0.147 |
| Lib Dem | -0.551 | -0.493 | -0.553 | -0.489 | -0.477 | -0.356 | -0.355 | 0.232 |
| Brexit Party | 0.546 | . | 0.490 | . | 0.469 | . | 0.399 | . |
| Reform UK | . | 0.277 | . | 0.187 | . | 0.614 | . | 0.580 |
| Vote | -0.776 | -0.887 | -0.790 | -0.801 | -0.532 | -0.477 | -0.336 | -0.286 |
| N | 532 | 518 | 532 | 518 | 532 | 542 | 532 | 542 |

**Figure 1:**
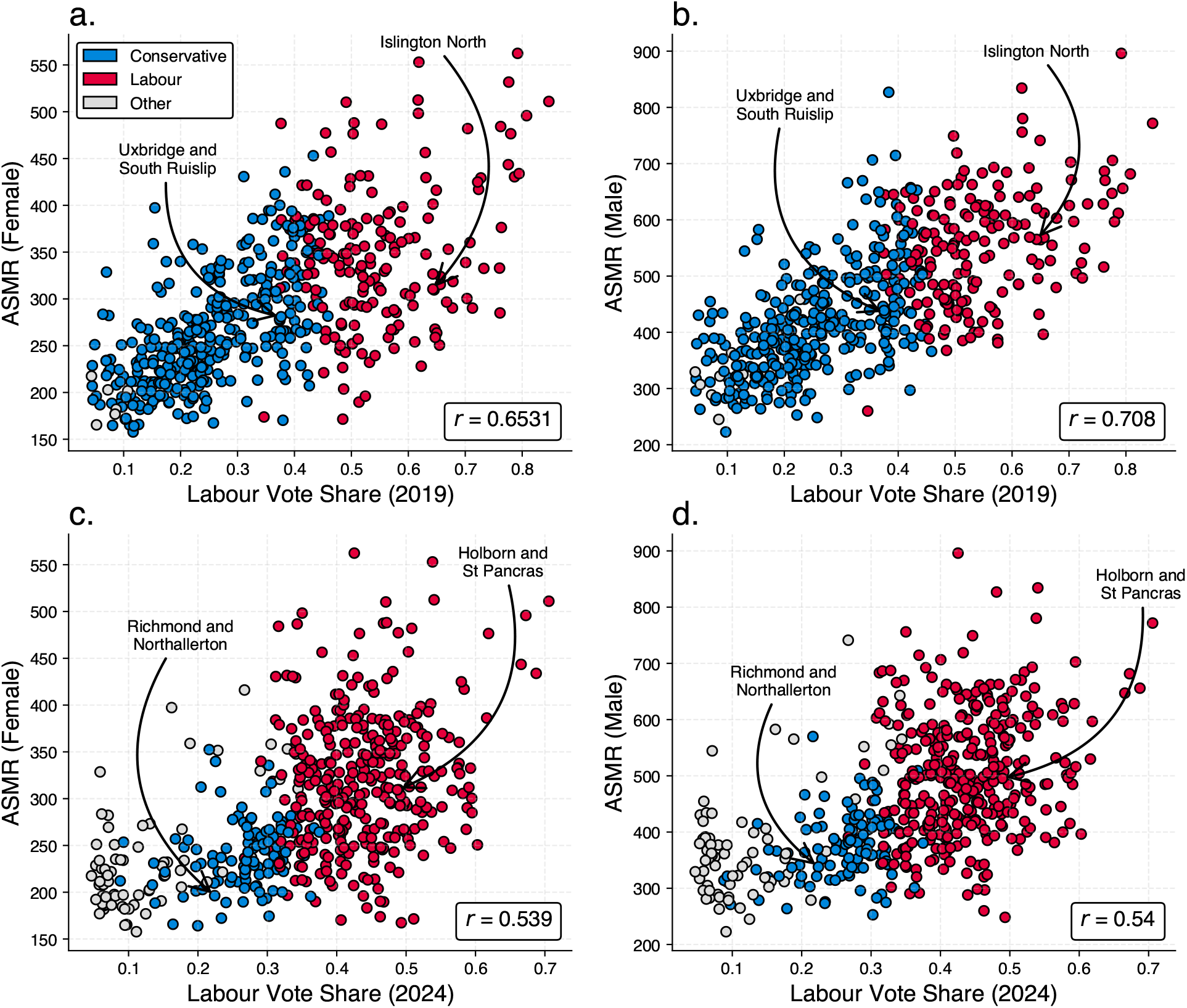
Correlations between voting behaviour and age standarised mortality at the constituency level in 2019 and 2024. Subfigures ‘a.’ and ‘b.’ show the correlation between Labour vote share across constituencies in 2019 and subfigures ‘c.’ and ‘d.’ in 2024 by Age Standardised Mortality Rates (ASMR) in 2021. Figures highlight the political party leader constituencies in 2019 (Conservative, Boris Johson: Uxbridge and South Ruislip; Labour, Jeremy Corbyn: Islington North) and 2024 (Conservative, Rishi Sunak: Richmond and Northallerton; Kier Starmer: Holborn and St. Pancras). Vote share data comes from the House of Commons Library, and mortality data from the Office for National Statistics. *r* indicates Pearson’s correlation coefficient (all p*<*0.0001). Supplementary material visualises the correlations between other party vote shares.

### 2.2 Methods

The relationship between voting patterns and ASMR, health, and disability by constituency was assessed using Pearson’s correlation coefficient (*r*). In the ASMR analysis, this was calculated for males and females separately (Figure 1). Bivariate choropleths were used to display the relationship between voting patterns and health and disability (Figure 2), with the correlations from that appearing in Table 1 (which contains all results).

**Figure 2:**
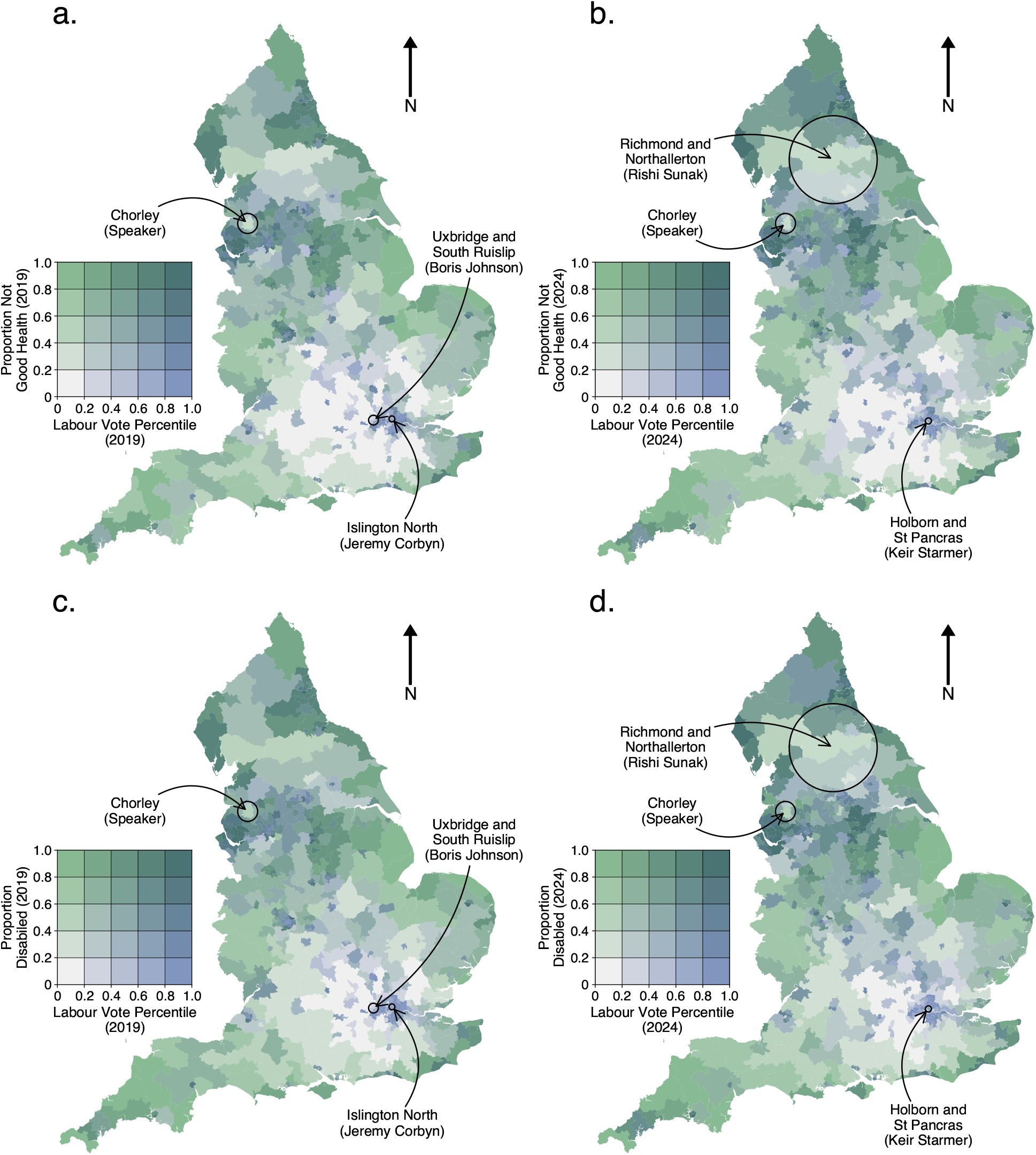
Bivariate choropleth map showing relationship between the proportion of the population with not good health and having a disability by Labour vote percentile in 2019 and 2024. Subfigures ‘a.’ and ‘b.’ show the proportion of people in not good health by Labour vote percentile in the 2019 (‘a.’) and 2024 (‘b.’) elections. Subfigures ‘c.’ and ‘d.’ show the proportion of people with a reported disability by Labour vote percentile in the elections in 2019 (‘c.’) and 2024 (‘d.’). Figures highlight the Speaker (Chorley) and political party leader constituencies in 2019 (Conservative, Boris Johson: Uxbridge and South Ruislip; Labour, Jeremy Corbyn: Islington North) and 2024 (Conservative, Rishi Sunak: Richmond and Northallerton; Kier Starmer: Holborn and St. Pancras). Vote share data comes from the House of Commons Library, and health and disability data comes from the Office for National Statistics. Disability is as defined in the main text.

## 3 Results

Figure 1 illustrates the contemporary relationship between voting patterns and mortality based on age-standardised mortality rate (ASMR) across the 532 constituencies contested in England in the 2019 and 2024 general elections The average ASMR across all 2019 constituency boundaries was 291 deaths per 100,000 for females and 459 for males, where higher values represent greater age standardised mortality. The mean values for constituencies that elected a Labour representative (n=179) in 2019 were 554.1 and 345.7 for males and females respectively, compared to 413.5 and 264.9 for Conservative constituencies (n=345). This was 504.04 (males) and 319.98 (females) for Labour, and 374.8 and 238.7 for males and females for the constituencies that elected a Conservative candidate in 2024. The ASMR for Richmond (Yorks) – Rishi Sunak’s constituency within the 2019 boundaries as the incumbent leader of the Conservative Party in the 2024 election – was 196.3 (95% CI 159.9-232.7) for females, and 338 (95% CI 289.5-386.6) for males in 2021. In comparison, Kier Stammer’s constituency, as the leader of the Labour Party and incumbent in Holborn and St Pancras, had an ASMR of 309.6 (95% CI 256.6-362.5) and 495.4 (95% CI 425.9-564.9) for females and males respectively. When observing the proportion of the vote share for Labour at the constituency level, there was a strong correlation with higher ASMR for males (*r*=0.708, *r*=0.54) and females (*r*=0.653, *r*=0.539) for 2019 and 2024 respectively. Results for other parties are available in our online supplementary material.

Figure 2 provides bivariate choropleth maps which display the relationship between the proportion of people with bad health (‘a.’, ‘b.’) and with a disability (‘c.’, ‘d.’) by Labour Party vote percentile in 2019 (subfigures ‘a.’, ‘c.’) and 2024 (‘b.’, ‘d.’). These maps highlight the relationship of ‘not good’ health and disability with Labour Party vote share across different geographical locations. Each colour on the map represents the strength of the association between variables. Labour vote share is correlated with health (subfigures ‘a.’, ‘b.’), but much less so with disability (Figures ‘c.’, ‘d.’). Considering Figure 2b., for instance, where we see a strong association (darker colour) in the constituency of Whitehaven and Workington, west of Sunak’s Richmond and Northallerton. This means that people within that area have both a higher proportion of health which is not good, and the constituency has a high vote share for Labour. These maps thus illustrate the geographical patterning of voting behaviour by health and disability. Table 1 provides a summary of the results. We see a generally strong negative correlation between ASMR for both sexes across both election periods for the Conservative and Liberal Democrat parties, as opposed to positive correlations for Labour, the Brexit Party (2019) and Reform UK (2024). An interesting finding is that while in 2024 we observe a negative relationship between Conservative voting and health and disability, this was positive in 2019. For the Liberal Democrats, the relationship was large and negative in both 2019 and 2024. Perhaps the most interesting result of all is that the strongest correlations are generally not observed with any specific party, but with the proportion of people who actually cast a vote.

## 4 Discussion

Although the goal of reducing geographical health inequalities was broadly similar amongst the governing Conservative and Labour parties over previous decades, we show a persistence in how voting behaviour is correlated with localised measures of mortality, health, and disability. Our results indicate a remarkable persistence in the patterning of voting behaviour by local health indicators, first demonstrated by Smith and Dorling, although with some striking differences between the 2019 and 2024 election. In line with previous research, a higher proportion of Labour voters were concentrated in geographical areas that had higher age-standardised mortality rates, and higher proportions of not good health and disability in both the 2019 and 2024 elections. There was a reduction in the correlation between votes cast for Labour in areas of higher mortality in the 2024 election, with correlation 0.54 for males and females 0.539. This is both lower than 2019 and lower than the previous estimates from elections in 1980s and 1990s.[8] This may however be a function of the fact that we are using the ‘closest’ constituencies to the original boundaries for which ASMR data is available.

In comparison to other countries, the correlation between voting patterns and measures of health inequalities such as ASMR are much stronger in England. For example, similar analysis finds no significant relationship between voting patterns and mortality in Australia. Counties that voted Republican in the 2016 US elections had overall worse health outcomes than those that voted Democrat; however, only a 2% decrease in median life expectancy was observed. A key factor behind the strong association between voting intensions and mortality is that, except for the 2019 ‘Get Brexit Done’ election, voting patterns are generally strongly correlated with measures of deprivation. In 2019, there were small positive correlations between Conservative Party vote share and measures of health and deprivation. This reflected a marked shift in the electoral and communication strategies of the Conservative party in 2019. Using the British Election Study, research has shown that Boris Johnson, the then Conservative Party leader at the time, used messaging that was targeted at the ‘red wall’ (traditionally Labour); areas termed the economically depressed ‘geographies of discontent’.[11] Similarly, the Brexit Party (2019) and Reform UK (2024) vote shares are both positively correlated with ASMR and with proportion of people reporting worse health and more disability. The findings show that although multiple governments aim to reduce health inequalities or ‘level up’ regions in England, inequalities are reflected in geographical voting patterns.

## 5 Strengths and Limitations

We go beyond previous work by not only examining sex stratified correlations between vote share for major parties and age standardised mortality rates, but we also provide additional insights from correlations with additional measures from the Census such as reported health and disability. Although we have extended our knowledge, the study also has several limitations. First, as it is based on official statistics, there are differences between the health inequality measures used in this study (i.e., ASMR) and those reported in a previous BMJ study (i.e., standardized mortality ratios for all-cause mortality). It is possible to explore other measures by electorate which have been shown to be associated with deprivation and health inequality.[12] Second, we are limited in the 2024 analysis of ASMR as a function of the fact that we need to map new and redrawn constituencies to their closest constituency as they were 2019. Third, we acknowledge that we are not using individual-level data that links voting behaviour with regional circumstances. Rather, we are using aggregated measures to look at the relationships between the proportion of people in a specific region who voted for a particular party with the concentration of mortality, health, and disability. This can risk what is termed an ‘ecological fallacy’; an event which occur when incorrect conclusions about individuals are drawn from data collected from data collected at the group or regional level. We also note that correlations do not provide evidence for causation between voting behaviour, mortality, health, or disability

## 6 Conclusions

Despite numerous efforts, health inequalities as measured by mortality, poor health and disabilities have remained, and, in some cases, increased in England. Although the relationship between mortality, poor health and disability with voting and politics is not a straightforward one, we once again find an enduring relationship between voting patterns and health inequalities at a constituency level.

## Data Availability

Replication materials are provided in full at:
https://github.com/crahal/voting_mortality_inequality

## Notes

### Competing Interest Statement

M.C.M. is a Trustee of the UK Biobank, on the Scientific Advisory Boards of Our Future Health and the LifeLines Biobank, and on the data management and advisory board of the Health and Retirement Survey.

### Funding Statement

Funding is gratefully acknowledged from the Health Foundation (REAL Demand Unit), the Leverhulme Trust (Grant RC-2018-003) for the Leverhulme Centre for Demographic Science, Nuffield College, and from the Economics and Social Research Council and MapIneq (101061645)

### Summary of Updates

New target variables, updated to 2024 election, with auxillery information (SII, HLE) removed.

## References

[1] Ian Holdroyd, Alice Vodden, Akash Srinivasan, Isla Kuhn, Clare Bambra, and John Alexander Ford. Systematic review of the effectiveness of the health inequalities strategy in england between 1999 and 2010. BMJ open, 12(9):e063137, 2022.

[2] Ben Barr, James Higgerson, and Margaret Whitehead. Investigating the impact of the english health inequalities strategy: time trend analysis. BMJ, 358, 2017.

[3] Donald Acheson et al. Independent inquiry into inequalities in health report. 2001.

[4] Mary Shaw, George Davey Smith, and Danny Dorling. Health inequalities and New Labour: how the promises compare with real progress. Bmj, 330(7498):1016–1021, 2005.

[5] Beata Coffey. Tackling health inequalities in england: A selected chronology up to august 2022. Royal Society of Medicine, pages 1–7, 2022.

[6] UK Parliament. Government action on major conditions and diseases: Statement made on 24 january 2023, 2023.

[7] Chris Ham. What needs to be done to make the nhs fit for the future?, 2022.

[8] George Davey Smith and Daniel Dorling. “I’m all right, John”: Voting patterns and mortality in England and Wales, 1981–92. Bmj, 313(7072):1573–1577, 1996.

[9] Office for National Statistics. Numbers and age-standardised mortality rates for premature deaths by parliamentary constituency and sex in 2021. https://www.ons.gov.uk/, 2021. [Online; accessed 9-February-2025].

[10] gov.uk. Definition of disability under the Equality Act of 2010. https://www.gov.uk/definition-of-disability-under-equality-act-2010, 2010. [Online; accessed 01-February-2025].

[11] Luke Cooper and Christabel Cooper. ‘get brexit done’: The new political divides of england and wales at the 2019 election. The Political Quarterly, 91(4):751–761, 2020.

[12] Xuejie Ding, Evelina T Akimova, Bo Zhao, Kasimir Dederichs, and Melinda C Mills. Prepayment meters strongly associated with multiple types of deprivation and emergency respiratory hospital admissions: an observational, cross-sectional study. J Epidemiol Community Health, 78(1):54–60, 2024.

